# Network Graph Representation of COVID-19 Scientific Publications to Aid Knowledge Discovery

**DOI:** 10.1101/2020.10.12.20211342

**Authors:** George Cernile, Trevor Heritage, Neil J Sebire, Ben Gordon, Taralyn Schwering, Shana Kazemlou, Yulia Borecki

**Author notes:** **Correspondence** Prof NJ Sebire, HDRUK, Welcome Trust, Euston Road, London UK.

## Abstract

**Introduction:** Numerous scientific journal articles have been rapidly published related to COVID-19 making navigation and understanding of relationships difficult.

**Methods:** A graph network was constructed from the publicly available CORD-19 database of COVID-19-related publications using an engine leveraging medical knowledgebases to identify discrete medical concepts and an open source tool (Gephi) used to visualise the network.

**Results:** The network shows connections between disease, medication and procedures identified from title and abstracts of 195,958 COVID-19 related publications (CORD-19 Dataset). Connections between terms with few publications, those unconnected to the main network and those irrelevant were not displayed. Nodes were coloured by knowledgebase and node size related to the number of publications containing the term. The dataset and visualisations made publicly accessible via a webtool.

**Conclusion:** Knowledge management approaches (text mining and graph networks) can effectively allow rapid navigation and exploration of entity interrelationships to improve understanding of diseases such as COVID-19.

## Introduction

There is urgency to accelerate research that can help contain the spread of the COVID-19 epidemic; to ensure that those affected are promptly diagnosed and receive optimal care, and to support research priorities in a way that leads to the development of global research platforms in preparation for the next disease epidemic; thus, allowing for accelerated research, R&D for diagnostics, therapeutics and vaccines and their timely access. In view of the urgency of this outbreak, the international community is mobilising to find ways to significantly accelerate the development of interventions.[1] Experts have identified key knowledge gaps and research priorities and shared scientific data on ongoing research, thereby accelerating the generation of critical scientific information to contribute to control the COVID 19 emergency.[2]

However, the pace and volume of research means that it is hard to stay up to date with the growing body of new scientific papers about the disease and the novel coronavirus that causes it. To mitigate this, many organisations are hosting digital collections holding thousands of freely available papers that can help researchers quickly find the information they seek. By one estimate, the COVID-19 literature published since January has reached more than 200,000 papers and is doubling every 30 days; among the biggest episodes of disease specific publication of scientific literature ever.[3]

One approach to navigating and searching such knowledge collections is through graph databases, which represent the connections between the semantic concepts with nodes, edges and other properties of the data. This allows semantic queries to search across the dataset to find relationships between papers on any set of data points. Such a graph displayed in a visualization tool gives an interactive overview of the nodes and connections between the concepts across the papers and allows one to move around and focus on what is interesting to the researcher.[4]

The aim of this short report is to demonstrate the feasibility of using a network graph approach for rapid navigation of the COVID-19 literature in a publicly available format and to present an openly available tool for exploring a COVID-19 knowledge dataset.

## Methods

The CORD-19 data set is a rapidly increasing open source collection of scholarly articles related to the corona virus.[5] As of the date August 8, 2020, the dataset consists of 207,311 papers from over 16,0000 sources. Articles available include title, abstract, authors, source, publication date, and in some cases full text.[6]

We used a proprietary natural language processing (NLP) and artificial intelligence (AI) engine that leverages the approach of heuristic segmentation (a fast heuristic search algorithm) and a knowledge-driven approach for concept identification, context determination, inferencing and extraction of corresponding values and units. The engine works with domain specific knowledgebases of clinical terms, concepts and rules that are tailored to the data to be extracted.[7]

In this study, we used a collection of ten knowledgebases consisting of a core knowledgebase and nine domain specific knowledgebases that were built using UMLS terms and updated with recently added terms specific to COVID-19: Core Oncology knowledgebase,Pharmacological substance (medications) (T121), Virus (T005), Therapeutic or preventative procedure (T061). Sign or symptom (T184), Disease or syndrome (T047), Gene or genome (T028), Immunologic factor (T129), Finding (T033), Body part, organ or organ component (T023).[8]

The title and abstract sections of all papers in the CORD-19 dataset were processed against the various knowledge sources to extract discrete data from each paper and stored in a database. Along with the discrete data, the following metadata was also stored: Cord uid (the unique identifier), Title, Abstract, Body text, Publication date, URL, Authors, Journal, KB – which of the 10 available knowledge sources was used to extract this term, Term category or question; ie: medication, virus, symptom, Paper Id – id of the paper in the CORD-19 dataset from which the term was extracted, Source section – either title or abstract. Generic terms were determined to have little significance for example, “Air”, “Water”, “Virus” and these were removed from the set of extracted concepts.

Networks created with the entire set of results and all the knowledge sources are very large with too many terms to visualize details in the data. For this reason, a subset of the data was selected to enable meaningful visual exploration by selecting a subset of the knowledge sources, paper sections and publication year for each network based on specific medical themes, e.g. treatments, cardiology etc. Duplicate terms (same term found in multiple knowledge sources) were consolidated to remove redundant data. For example, “obesity” is included in both the “Symptoms and Side Effects” and the “Disease or Syndrome” knowledge sources.

For each term found in a paper, a link was created to every other term in the same paper. The culmination of these links for all papers resulted in the network structure where the weight of a connection between any two terms was determined by the number of papers linking the terms. Additional filtering was performed to refine the scope of the network and removal of noise; links with low weights were removed, as were links with terms that were disconnected from the rest of the network.

The open-source software tool Gephi was used to create a visualisation of the network using the collections of terms and connections that made up the network structure.[9] Network nodes were coloured based on the knowledge source, node sizes are proportional to the frequency of each term, the connection weight (edge thickness) was based on the number of associated papers. The networks were exported and visualised in an HTML website using the Sigma JS JavaScript library.

## Results

A total of 207,311 publications from the CORD-19 dataset were processed using the NLP engine. In total 3,357,328 total entities were extracted from 195,958 of these papers consisting of 44,494 unique terms. Four network graphs were generated using this extracted data: Cardiological Diseases, Lung Diseases, Title Network, Treatment Network.(Figure; https://nlp.inspirata.com/networkvisualisations/treatmentnetwork/#). The filters applied to create each of the networks and the number of terms, edges and papers involved in each network is displayed in the Table.

**Figure.**
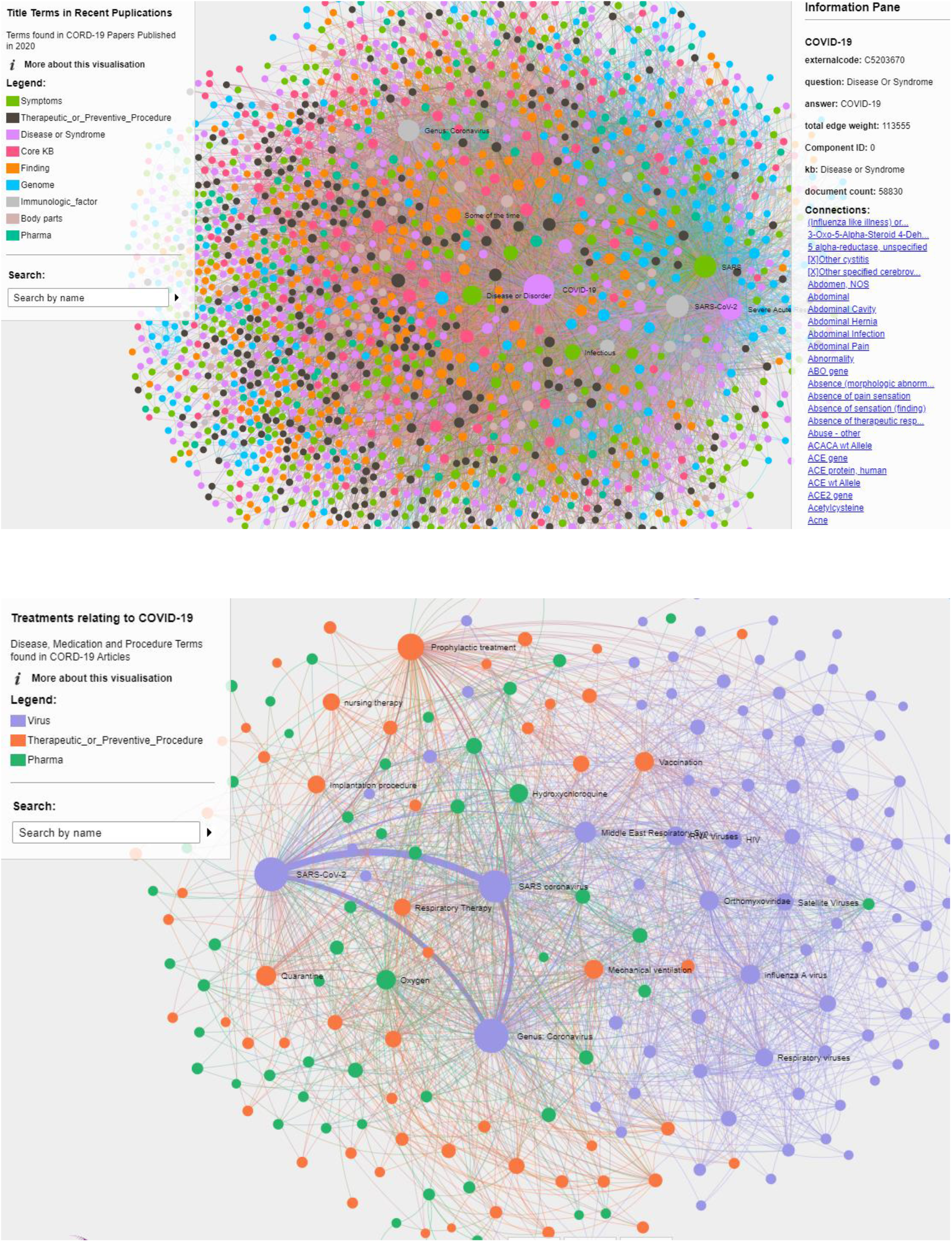
Example network graphs including High density network showing all concepts associated with COVID-19 (Top) and treatment map for COVID-19 (Bottom)

**Table 1:**
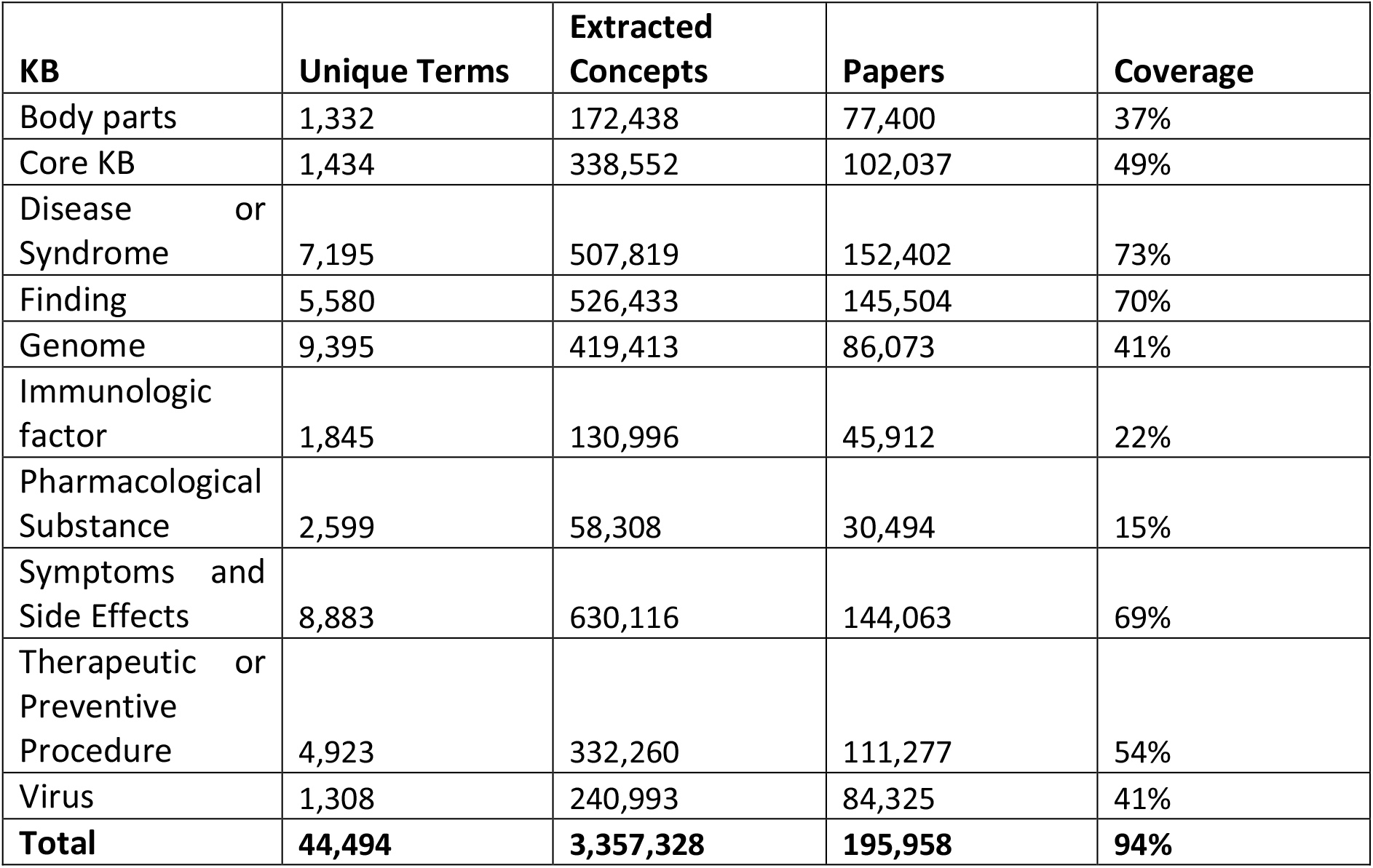
Extracted concepts from CORD-19 dataset by Knowledgebase (semantic type) showing the number of unique terms found and the total number of extracted values from each knowledge base, as well as the number of papers containing terms from that knowledge base, and percentage coverage across the entire dataset.

**Table 2:** Filters applied to four networks with links to visualizations showing the filters and the number of terms, edges and papers involved in each network.

|  | **Title Network** | **Treatment Network** | **Lung Diseases** | **Cardio Diseases** |
| --- | --- | --- | --- | --- |
| **Knowledge Baes** | ALL | Therapeutic or prevention procedure; Pharmacological Substances; Virus | Symptoms and Side Effects; Disease or Syndrome; Virus | Symptoms and Side Effects; Disease or Syndrome; Virus |
| **Sections** | Title | Title; Abstract | Title; Abstract | Title; Abstract |
| **Publication Years** | 2020 | 2015-2020 | 2015-2020 | 2015-2020 |
| **Edge Weight cutoff** | 5 | 5 | 10 | 25 |
| **Manual Filtering** | no | yes | yes | yes |
| **# papers** | 34,032 | 36,801 | 52,258 | 35,282 |
| **# nodes (terms)** | 2,586 | 206 | 86 | 49 |
| **# edges (connections)** | 16,735 | 1,739 | 994 | 277 |
| **Link** | https://nlp.inspirata.com/NetworkVisualisations/TitleNetwork/ | https://nlp.inspirata.com/NetworkVisualisations/TreatmentNetwork/ | https://nlp.inspirata.com/NetworkVisualisations/LungNetwork/ | https://nlp.inspirata.com/NetworkVisualisations/CardioNetwork/ |

## Discussion

Recently there have been several initiatives to explore knowledge graphs in medical data and with some applied to aspects of COVID-19 associated published literature.[10,11] This study has demonstrated the feasibility of using a graph database approach to create a targeted concept association networks as an interactive way to allow users to easily navigate the rapidly growing COVID-19 related literature, and particularly as a way of understanding and exploring the relationships between the key concepts within this corpus of literature articles.

This approach is applicable to any collection of scientific literature such as PubMed or Clinical Trials.gov, or proprietary document management systems. Specific lexical terms and knowledge sources can be utilized from the UMLS collection or other publicly available sources and imported for use with NLP/AI Engines.

One constraint is that the network size increases as more knowledge sources are added. As a consequence, methods for pruning the network to enable visual exploration are required. As the density of the network increased it becomes more difficult to interactively explore without a-priori knowledge of the specific knowledge sources. Another limitation is that the network only shows the first level connections or the direct connection between papers and concepts. It does not find connections between concepts that span several papers – although this can be achieved by traversing the network visually.

We addressed these limitations of network size and the search for deep connections by implementing a breadth-first search on the network structure. The search is efficient and can be applied across very large networks, even when all the knowledge sources are used simultaneously. This approach can find the shortest path connections (the trail of papers) between any concepts.

This study has demonstrated that an approach using graph databases and network analysis can be developed rapidly and is a useful approach for understanding large volumes of medical literature, quickly grasping the current state of our knowledge, and discovering previously unknown or unnoticed relationships between emerging medical concepts. The unusual circumstances of a global pandemic have given rise to assembly of an unprecedented volume of medical literature and this work demonstrates a powerful approach to condensing the literature into insights that help us fight this disease. Further development of this approach will enable ongoing analysis and deep searching of large collections of literature, such as PubMed, and application to other disease areas, as well as for target or biomarker discovery.[12–14]

## Data Availability

all data is publicly available through the links provided

https://nlp.inspirata.com/networkvisualisations/treatmentnetwork/#).

